# Cumulative In-Context Learning versus Simple Historical Weighting for Real-Time Geographic Origin Identification of Ongoing Epidemic Waves: A Comparative Evaluation Using Eight COVID-19 Waves in Japan

**DOI:** 10.64898/2026.05.23.26353936

**Authors:** Shin-ichi Nakagawa, Akira Yamamoto

**Affiliations:** Research Institute of Info-Communication Medicine (RinCOM); Faculty of Health Data Science, Juntendo University

**Author notes:** Corresponding author: Shin-ichi Nakagawa.

**Keywords:** In-context learning, Large language model, Epidemic origin identification, Cumulative learning, COVID-19, Surveillance

## Abstract

**Background:** Identifying the geographic origin of epidemic waves early is critical for targeted public health responses. Conventional statistical methods for wave origin estimation rely on fixed algorithms applied to case count time-series data and treat each wave independently. Large language models (LLMs) offer a novel alternative through cumulative learning—the ability to incorporate confirmed epidemiological findings from prior waves into predictions for subsequent waves. Whether this approach outperforms conventional statistical baselines in early detection, and whether the same cumulative learning principle can be implemented in transparent statistical methods, remains unknown.

**Methods:** We compared three computational approaches across eight COVID-19 epidemic waves in Japan (Waves 2–8, 2020–2023): (1) non-cumulative statistical baselines (B0–B5) treating each wave independently; (2) a cumulative-learning LLM (Claude Haiku) receiving confirmed origins from all prior waves as in-context historical knowledge; and (3) cumulative calculation statistical baselines implementing the identical historical weighting mechanism as a transparent arithmetic score. We additionally evaluated a non-cumulative LLM condition—receiving only current-wave data—to isolate the contribution of intrinsic LLM geographic reasoning from accumulated historical knowledge. All approaches were evaluated at 7, 14, 21, and 28 days after wave onset and validated against genomically confirmed wave origins.

**Results:** Cumulative calculation statistical baselines (B1, B3) achieved mean F1 = 0.51 at 14 days after wave onset, performing comparably to the cumulative-learning LLM (F1 = 0.52) and outperforming all non-cumulative statistical baselines (F1 = 0.41–0.46). Wave 7 (Omicron BA.5) was correctly identified at 14 days by both methods (F1 = 1.00). Wave 6 (Omicron BA.1) was undetectable by all methods (F1 = 0.00), consistent with an origin outside the domestic surveillance system.

**Conclusions:** The cumulative historical weighting mechanism—not LLM reasoning per se—drives performance improvement, as transparent arithmetic implementation matches LLM accuracy. However, the non-cumulative LLM achieves F1 = 0.46 without any historical context, suggesting substantial intrinsic geographic reasoning capacity. These findings advance understanding of when and why in-context learning confers advantage, and provide a deployable, spreadsheet-implementable method for real-time epidemic origin identification requiring no AI infrastructure.

## 1. Introduction

Rapid identification of the geographic origin of epidemic waves is a fundamental challenge in infectious disease surveillance. Knowing where a new wave begins allows public health authorities to implement targeted interventions—contact tracing, enhanced surveillance, travel advisories—before widespread community transmission occurs [1]. In large, geographically diverse countries such as Japan, where epidemic dynamics vary substantially by region, early origin identification could substantially improve the timeliness and efficiency of public health responses. The practical value of early origin identification lies in its ability to concentrate limited surveillance and response resources on candidate regions before the wave spreads nationally, potentially shortening the interval between first emergence and effective public health action. However, reliable origin identification typically relies on genomic surveillance, which requires specialized laboratory infrastructure, considerable time from sample collection to result, and financial resources that are not universally available [2]. In routine surveillance settings, genomic confirmation of wave origins often lags weeks behind clinical case detection, limiting its operational value for real-time decision-making. A system capable of providing probabilistic origin estimates from routinely available case count data—without waiting for genomic confirmation—would represent a meaningful advance in epidemic preparedness.

Conventional data-driven approaches to epidemic origin estimation include cross-correlation analysis, Granger causality testing, early growth rate metrics, and early onset day identification [3,4]. These methods analyze prefecture-level or regional case count time-series to infer which areas showed the earliest and most rapid increases in incidence. While computationally tractable and requiring only routinely available surveillance data, they share a fundamental limitation: they treat each wave in isolation, without incorporating knowledge accumulated from prior waves about which regions have historically served as epidemic origins, or about the immune landscape shaped by past infection. This wave-independent design means that useful epidemiological knowledge—for example, that certain coastal or metropolitan regions have repeatedly served as first points of variant introduction—cannot be leveraged to improve predictions for subsequent waves. Addressing this limitation requires an approach capable of integrating and applying historical patterns across waves in a systematic and scalable way.

Large language models (LLMs) have recently seen rapid expansion in epidemiological applications. Existing studies have focused primarily on forecasting case counts or estimating epidemic scale: Du et al. proposed a framework integrating genomic surveillance data and public health policy for short-term forecasting of COVID-19 hospitalizations using LLMs [6], and Jin et al. demonstrated a general-purpose approach for applying LLMs to time-series forecasting [8]. However, none of these approaches have as their primary aim the identification of the geographic origin of epidemic waves—that is, where a wave begins. The key advantage of LLMs for this application lies in their ability to incorporate confirmed epidemiological findings from prior waves into the estimation of the next wave. By providing the LLM with past information—variant designations, confirmed origin regions, and regional cumulative infection patterns—before analyzing a new wave, contextual reasoning becomes possible that is unavailable to conventional statistical methods that treat each wave independently, enabling real-time origin prediction with potentially higher accuracy than conventional approaches.

No prior study has systematically evaluated whether a general-purpose LLM, applied without fine-tuning or architectural modification, can identify the geographic origins of epidemic waves from routinely available case count data, or how its performance compares to established statistical baselines under identical early-detection conditions. Furthermore, the question of whether the cumulative-learning advantage of LLMs can be replicated using transparent, interpretable statistical methods has not been addressed.

In this study, we compared six conventional statistical methods (B0–B5) against a cumulative-learning LLM approach across eight COVID-19 epidemic waves in Japan, using data from the first 7 to 28 days after wave onset. We further evaluated cumulative-learning versions of the statistical baselines, in which confirmed genomic origins from prior waves are incorporated as a weighted bonus term. Japan provides an ideal setting for this evaluation: it experienced eight well-characterized epidemic waves spanning multiple variants over a three-year period, maintains publicly available prefecture-level surveillance data through the Ministry of Health, Labour and Welfare open data portal, and has documented genomic confirmation of wave origins from the National Institute of Infectious Diseases and prefectural health authorities that can serve as ground truth for validation. The sequential nature of these waves—each building on the epidemiological history of prior waves—makes Japan’s COVID-19 experience a uniquely informative testbed for evaluating cumulative-learning approaches to epidemic origin identification.

## 2. Methods

### 2.1 Data Sources

Daily prefecture-level COVID-19 confirmed case counts were obtained from the Ministry of Health, Labour and Welfare (MHLW) open data portal (https://www.mhlw.go.jp/stf/covid-19/open-data.html). Data spanned January 2020 through May 2023, encompassing eight epidemic waves across four major variant periods. Population-adjusted rates (cases per 100,000 population) were calculated using prefecture-level population estimates from the Statistics Bureau of Japan. All data underwent 7-day centered moving average smoothing to reduce day-of-week reporting artifacts before analysis.

### 2.2 Regional Block Definition

Origin identification at the individual prefecture level creates a sparse event problem: with 47 prefectures and typically only 2–4 confirmed origin regions per wave, F1-based performance evaluation becomes unstable. To ensure a practically tractable analysis, Japan’s 47 prefectures were therefore aggregated into 11 regional blocks based on geographic proximity and inter-prefectural epidemiological characteristics. This aggregation is also consistent with public health practice, in which interventions such as enhanced contact tracing, travel advisories, and reallocation of surveillance resources are typically implemented at a regional rather than individual prefecture level. The 11 regional blocks were defined as follows: Hokkaido, Tohoku (6 prefectures: Aomori, Iwate, Miyagi, Akita, Yamagata, Fukushima), Kanto (7 prefectures: Ibaraki, Tochigi, Gunma, Saitama, Chiba, Tokyo, Kanagawa), Koshin-etsu (3 prefectures: Yamanashi, Nagano, Niigata), Hokuriku (3 prefectures: Toyama, Ishikawa, Fukui), Tokai (4 prefectures: Aichi, Gifu, Shizuoka, Mie), Kinki (6 prefectures: Shiga, Kyoto, Osaka, Hyogo, Nara, Wakayama), Chugoku (5 prefectures: Tottori, Shimane, Okayama, Hiroshima, Yamaguchi), Shikoku (4 prefectures: Tokushima, Kagawa, Ehime, Kochi), Kyushu (7 prefectures: Fukuoka, Saga, Nagasaki, Kumamoto, Oita, Miyazaki, Kagoshima), and Okinawa. Regional case rates were calculated as population-weighted averages of constituent prefecture rates using population estimates from the Statistics Bureau of Japan.

### 2.3 Wave Definition and Genomic Validation

Eight epidemic waves were defined based on national case count dynamics and variant genomic surveillance reports from the National Institute of Infectious Diseases (NIID) and the Ministry of Health, Labour and Welfare. Wave boundaries were established at the dates of sustained national incidence transitions. Wave origins were validated against genomically confirmed data from peer-reviewed surveillance studies, NIID reports, and official prefectural health authority announcements. Evidence quality was rated as high (confirmation by two or more independent genomic sources including peer-reviewed literature or official NIID reports) or moderate (single genomic source or primary epidemiological linkage). Genomically confirmed origins for each wave were as follows: Wave 1 (Ancestral, March–June 2020): Kanto, Kinki, Hokkaido (high confidence); Wave 2 (Ancestral, June–October 2020): Kanto, Okinawa, Tokai, Kyushu (high confidence); Wave 3 (Ancestral/Alpha, October 2020–February 2021): Hokkaido, Kanto, Kinki (high confidence); Wave 4 (Alpha, March–June 2021): Kinki, Tohoku (high confidence); Wave 5 (Delta, June–November 2021): Okinawa, Kanto (high confidence); Wave 6 (Omicron BA.1, November 2021–June 2022): Okinawa, Chugoku (moderate confidence); Wave 7 (Omicron BA.5, July–October 2022): Kanto, Okinawa, Kinki (high confidence); Wave 8 (BA.5/XBB, October 2022–February 2023): Hokkaido, Kanto (high confidence). Wave 6 (Omicron BA.1) origins in Okinawa and Chugoku were confirmed through prefectural genomic analyses and official municipal reports consistent with early cluster events, though independent peer-reviewed confirmation from NIID genomic surveillance remains limited; results for this wave should be interpreted with caution.

### 2.4 Baseline Detection Methods

Six baseline methods (B0–B5) and a non-cumulative LLM condition were evaluated using only data from the first N days (N = 7, 14, 21, 28) after wave onset, treating each wave independently without incorporating knowledge from prior waves. No single hypothesis about what distinguishes an origin region is guaranteed to hold across all waves and variants; a parallel evaluation across all candidate methods maximizes the probability that the correct origin signal is captured. For each method, the top-3 ranked regions were designated as predicted origins. Cumulative extensions are described in Section 2.5.

#### B0 – Early Onset Day

The date on which a region first exceeds a minimum incidence threshold is a classical early warning indicator in infectious disease surveillance [20]. The rationale for origin detection is that the region where a wave begins should exceed the detection threshold before all others. For region r, the onset day is defined as: dL(r) = min{t: X_r(t) ≥ θ}, where X_r(t) is the population-adjusted daily incidence (cases per 100,000) and θ = 0.1/100,000 is the detection threshold, selected based on NIID surveillance practice for defining wave initiation in Japanese prefectural data [18]. Regions were ranked in ascending order of dL(r); regions not yet exceeding θ within the N-day window were assigned the lowest rank.

#### B1 – Peak Infection Rate

The maximum incidence attained during the early observation window reflects the intensity of local transmission. In a wave propagating outward from its geographic origin, the origin region is expected to accumulate the highest local intensity earliest [5,34]. The B1 score for region r is: B1(r, N) = max_{t ≤ N} X_r(t). Regions were ranked in descending order of B1(r, N). This metric is robust to day-of-week reporting artifacts after 7-day moving average smoothing and requires no parametric assumptions about epidemic trajectory.

#### B2 – OLS Growth Rate

The early exponential growth rate is a fundamental quantity in epidemic theory, capturing the speed of transmission expansion [4,34]. An origin region is expected to exhibit faster early growth than regions receiving the wave later. An ordinary least squares (OLS) regression was fit to the N-day incidence time series for each region: X_r(t) = β_r · t + α_r + ε_r(t). The normalized growth rate score is: B2(r, N) = [βL_r / XL_r − min_{r’} (βL_{r’} / XL_{r’})] / range_{r’} (βL_{r’} / XL_{r’}), where βL_r is the OLS slope and XL_r is the mean incidence over days 1 to N. Dividing by the mean removes the effect of absolute incidence level, isolating relative growth velocity. The score is min-max normalized to [0,1], and regions were ranked in descending order.

#### B3 – Cumulative Infection Rate

Cumulative incidence over the early observation window integrates both the intensity and duration of early transmission, providing a measure complementary to the instantaneous peak (B1). The origin region is expected to accumulate more total cases early in the wave [5]. The B3 score is: B3(r, N) = Σ_{t=1}^{N} X_r(t). Regions were ranked in descending order of B3(r, N). Unlike B1, this metric is less sensitive to single-day reporting spikes and captures sustained early elevation, making it appropriate when daily case counts are noisy.

#### B4 – Cross-correlation Lead Score

Spatial cross-correlation analysis identifies the temporal lead-lag structure between regional time series, quantifying which regions show incidence rises that systematically precede others—a pattern consistent with origin-to-destination wave propagation [16]. For each ordered pair of regions (r, r’), the lag maximizing the cross-correlation of de-meaned incidence series was identified. The lead score for region r is: B4(r) = count{r’ not equal r: optimal lag(r,r’) is negative} divided by (R-1), where R = 11 is the number of regional blocks. A higher score indicates that region r’s incidence series consistently precedes those of other regions, operationalizing the concept of epidemic spatial leadership.

#### B5 – Granger Causality Score

Granger causality testing [3] assesses whether past values of one time series improve prediction of another beyond autoregressive terms alone—operationalizing temporal predictive priority between regions. This approach has been applied to COVID-19 regional transmission dynamics to identify leading areas [17]. A bivariate Granger causality test with lag L = 3 days and second-order differencing for stationarity was applied to each ordered region pair (r, r’). The score for region r is: B5(r) = count{r’ not equal r: X_r Granger-causes X_r’ at p less than 0.05} divided by (R-1). A region with a high B5 score predicts future incidence rises in many other regions, suggesting it is a transmission source rather than a recipient of the wave.

### 2.5 Cumulative Calculation Methods

Epidemic waves do not occur in isolation. Each wave leaves a geographic footprint—certain regions consistently emerge as early foci across multiple waves—that carries predictive value for subsequent waves. This observation motivates a cumulative-learning framework, in which knowledge accumulated from prior waves is systematically incorporated when predicting the origin of the next wave. This approach is conceptually analogous to Bayesian updating: prior knowledge (historical origin patterns) is combined with current evidence (early incidence data) to form a posterior prediction. However, we do not specify a formal prior distribution; instead, historical origin frequencies serve as an empirical weight added to current-wave scores—a deliberate simplification that keeps every weighting decision transparent and auditable. Crucially, for Wave N, only data from Waves 1 through N−1 are used to construct the cumulative weight; Wave N itself contributes no information to its own prediction, eliminating data leakage and ensuring a fair comparison with non-cumulative methods. For the LLM component, cumulative learning takes a different form: historical origin information is provided as natural language context within the prompt, enabling in-context reasoning rather than parameter updating. The model’s weights remain unchanged throughout, so overfitting in the conventional sense does not apply.

#### 2.5.1 Cumulative Calculation: Statistical Baselines

To evaluate whether the cumulative calculation approach can reproduce the LLM’s advantage using transparent, interpretable methods, we implemented cumulative calculation versions of B1, B2, and B3. The complete procedure is summarized in Box 1. The cumulative score is defined as Score_cumul(r) = Score_baseline(r) + λ × P(r,n), with λ = 0.75 selected from a sensitivity analysis across λ ∈ {0.00, 0.25, 0.50, 0.75, 1.00}. The top-3 regions by Score_cumul were designated as predicted origins. The complete four-step implementation is given in Box 1. Baseline scores for B1–B5 are normalized to [0,1] as follows. B1:

B1(r,N) = max_{t≤N} X_r(t) / max_{r’} max_{t≤N} X_{r’}(t). B2:

B2(r,N) = [β□_r / X□_r − min(β□/X□)] / range(β□/X□), where β□_r is the OLS slope over days 1–N. B3: B3(r,N) = Σ_{t=1}^{N} X_r(t) / max_{r’} Σ_{t=1}^{N} X_{r’}(t). B4: cross-correlation lead score (proportion of regions for which r’s series leads at positive lag); B5: Granger causality score (proportion of regions significantly Granger-caused by r at p < 0.05, lag = 3). A sensitivity analysis across λ ∈ {0.00, 0.25, 0.50, 0.75, 1.00} confirmed that B1 and B3 are stable at λ ≥ 0.25 (Table S1). A leave-one-wave-out analysis confirmed that λ ≥ 0.25 was optimal in 6 of 7 waves; the exception was Wave 8 (BA.5/XBB), where successive same-lineage waves may reduce the predictive value of historical weighting (Table S2).

##### Box 1. Cumulative Calculation Method: Four-Step Implementation

**Given:** Wave n, Region r (r = 1–11), Confirmed origins G_1,…,G_{n−1} from prior waves

**Step 1 — Historical origin frequency (empirical prior):**

P(r,n) = 1/(n−1) × Σ_{k=1}^{n−1} I(r ∈ G_k)

*“How often has region r been a confirmed origin in past waves?” — accumulates automatically with each successive wave*

**Step 2 — Current-wave score from first N days (select one baseline):**

B1(r,N) = max_{t≤N} X_r(t) / max_{r’} max_{t≤N} X_{r’}(t) [peak rate, normalized 0–1]

**Step 3 — Combine (Bayesian-inspired, no distributional assumptions required):**

Score_cumul(r) = Score_baseline(r) + λ × P(r,n) (λ = 0.75)

**Step 4 — Select predicted origins:**

Top-3 regions by Score_cumul → predicted origins [implementable in any spreadsheet]

#### 2.5.2 Cumulative Learning: LLM

A general-purpose LLM (Claude Haiku, claude-haiku-4-5-20251001; Anthropic) was used without fine-tuning or architectural modification. API configuration: endpoint https://api.anthropic.com/v1/messages; max_tokens = 300; temperature = default (1.0). Output format: structured JSON. All API calls were stateless (no conversation history carried over between calls). For each target wave, the LLM received a structured prompt containing: (1) the infection rate time-series for the first N days aggregated to the 11 regional blocks as a ranked list of rates per 100,000 population, (2) the Early Onset Day indicator (B0) computed from the same N-day window, and (3) cumulative historical context—for each prior wave, the confirmed genomic origins, the regions with highest and lowest cumulative infection rates, and the variant designation. The LLM was instructed to identify the top-3 regional blocks most likely to be the wave origin, responding in structured JSON format. Three independent runs were conducted per wave-day combination, and mean F1 scores were reported.

The key feature of the prompt design is the integration of two information streams: current-wave early signals and accumulated historical knowledge. To illustrate, the prompt for a target wave with 14 days of data took the following form (abbreviated): “You are an epidemiologist. A new COVID-19 wave [target wave, variant] has just started in Japan. You only have data from the FIRST 14 DAYS of this wave. Japan has 11 regions: Hokkaido, Tohoku, Kanto, […all 11 regions…]. HISTORICAL confirmed wave data: --- Wave 1 (Ancestral) --- Earliest onset: Kanto(3days), Kinki(5days), Hokkaido(7days). Highest cumulative rate: Kanto(12.4), Kinki(8.1), Hokkaido(6.3). Confirmed origins: Kanto, Kinki, Hokkaido. […all prior waves…]. CURRENT WAVE first 14 days (per 100K): Okinawa: 2.31/100K, onset=3days, rising. Kanto: 1.87/100K, onset=5days, rising. […all 11 regions…]. Based on historical patterns AND early signals, identify the TOP 3 regions most likely to be the ORIGIN. Respond ONLY with JSON: {‘top3_regions’: [‘Region1’, ‘Region2’, ‘Region3’], ‘reasoning’: ‘brief’}.” This prompt structure encodes the cumulative-learning mechanism: by seeing which regions were confirmed origins in all prior waves, the LLM can apply historical pattern recognition to the current wave. The historical context grows with each successive wave, providing progressively richer prior information. The prompt for the cumulative-learning condition followed this structure (abbreviated): “You are an epidemiologist. A new COVID-19 wave ({wave_name}, {variant}) has started in Japan. You have data from the FIRST {N} DAYS only. […11 regional blocks listed…] EARLY ONSET DAY (first day exceeding 0.1/100K): {region}: day {onset_day} […all 11 regions…] CURRENT WAVE data (per 100K): {region}: {rate}/100K, onset day {day}, trend: rising/stable […all regions…] HISTORICAL confirmed wave data: --- Wave 1 (Ancestral) --- Earliest onset: {regions with days}. Highest cumulative rate: {regions with values}. Confirmed genomic origins: {confirmed_origins}. […all prior waves…] Based on historical patterns AND early signals, identify the TOP 3 regional blocks most likely to be the ORIGIN. Respond ONLY with JSON: {‘top3_regions’: [‘Region1’, ‘Region2’, ‘Region3’], ‘reasoning’: ‘brief’}.” For the non-cumulative condition, the HISTORICAL section was omitted entirely; the model received only current-wave data.

Each time-point prediction (7, 14, 21, and 28 days after wave onset) was conducted as a fully independent API call. The prediction result from a prior time point was not included in the input for subsequent time points; each call used only the case data accumulated up to that time point and the confirmed historical context from all prior waves. This design mirrors real-time operational use, in which the system would be queried at discrete time points using the most current available data, and ensures that performance differences across time points reflect the effect of data accumulation rather than cross-time-point information carry-over. This cumulative-learning design ensures that information from Wave k is incorporated for predicting Wave k+1, with no future information leakage. Wave 1 was used as historical context only and was not evaluated as a prediction target.

A general-purpose LLM (Claude Haiku, claude-haiku-4-5-20251001; Anthropic) was used without fine-tuning or architectural modification.

### 2.6 Evaluation Metric

Performance was assessed using the F1 score computed against genomically confirmed regional origins. For each wave-method-timepoint combination, Precision was defined as the proportion of predicted regions that were confirmed origins (tp / (tp + fp)); Recall as the proportion of confirmed origins that were predicted (tp / (tp + fn)); and F1 as their harmonic mean (2 × Precision × Recall / (Precision + Recall)). When no regions were predicted or no confirmed origins existed, F1 was set to 0. All methods predicted exactly 3 origin regions; confirmed origin counts ranged from 2 to 4 across waves. Mean F1 scores were calculated as unweighted averages across Waves 2–8 (7 waves); Wave 1 was excluded as it served as historical context only. Wave 6 results are reported separately given evidence for an origin outside the domestic surveillance system, and its inclusion in the mean is noted to avoid misleading interpretation of overall method performance.

## 3. Results

### 3.1 Performance of Statistical Baselines

At 14 days after wave onset, conventional statistical methods achieved mean F1 scores of 0.41 (B0, Early Onset Day), 0.46 (B1, peak infection rate), 0.32 (B2, OLS growth rate), 0.46 (B3, cumulative infection rate), 0.26 (B4, cross-correlation), and 0.31 (B5, Granger causality) (Figure 2, Figure 3). The non-cumulative LLM—receiving only current-wave data without any historical context—achieved mean F1 = 0.46 at 14 days, matching B1 and B3 and outperforming B0, B2, B4, and B5. Notably, the non-cumulative LLM detected Wave 6 (Omicron BA.1; F1 = 0.40), whereas all statistical baselines failed (F1 = 0.00), suggesting that LLM geographic reasoning captures signals beyond simple incidence metrics. B2, B4, and B5 performed poorly across all time points, reflecting the instability of growth rate estimation and causality testing with limited early-phase data of 7–14 days. B0, B1, and B3 showed more consistent performance, with B1 and B3 reaching mean F1 = 0.62 at 21–28 days when more data accumulated.

**Figure 1.**
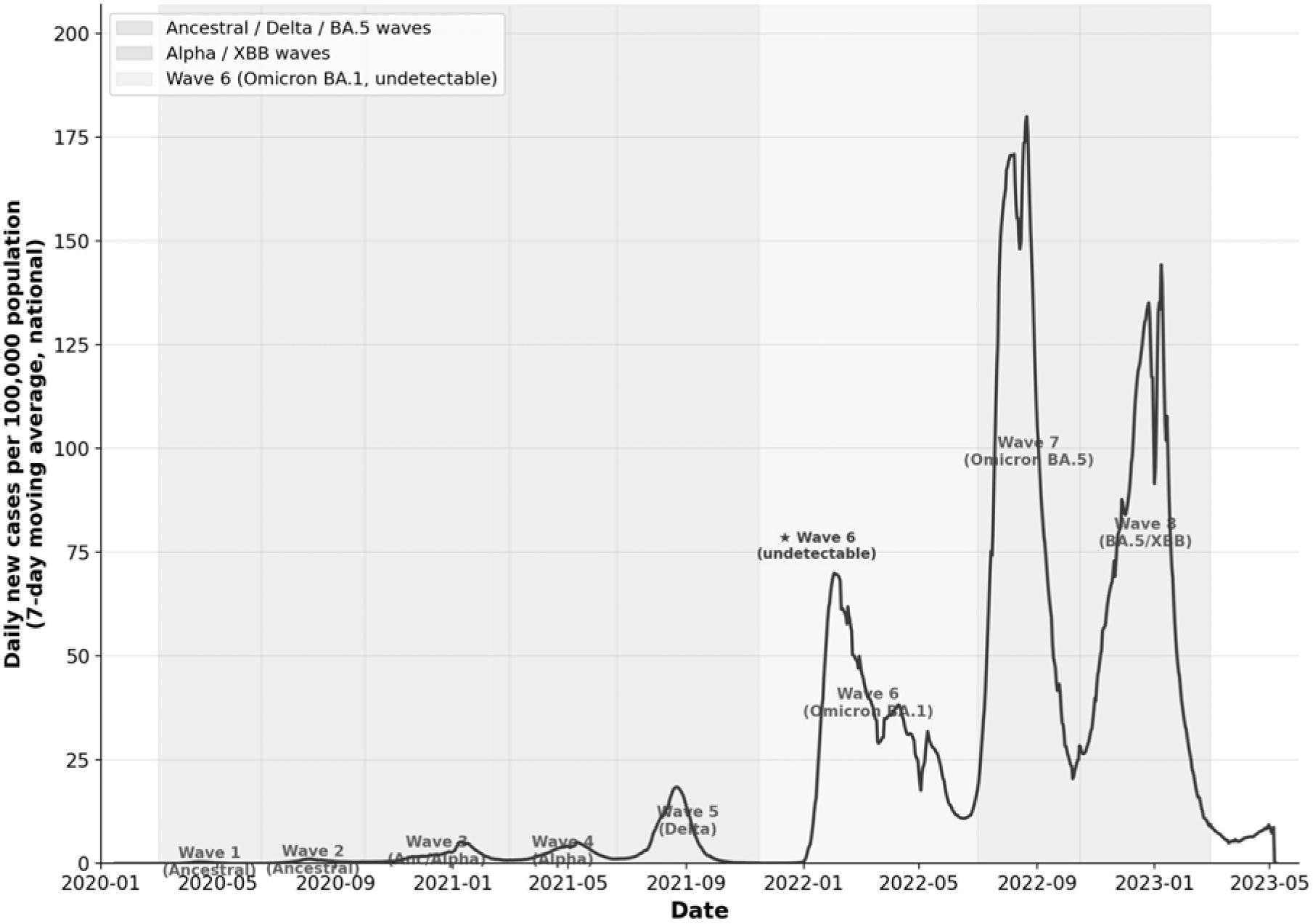
National COVID-19 epidemic trends in Japan across eight waves (March 2020–May 2023) Daily new COVID-19 cases per 100,000 population (7-day moving average, national level) across eight epidemic waves in Japan. Wave periods are shaded by variant group: blue (Ancestral/Delta/BA.5 waves), green (Alpha/XBB waves), and pink (Wave 6, Omicron BA.1). Wave 6 is marked with a red star to indicate that it was undetectable by all methods evaluated in this study (F1 = 0.00 at all time points). Wave labels and variant designations are shown within each wave period.

**Figure 2.**
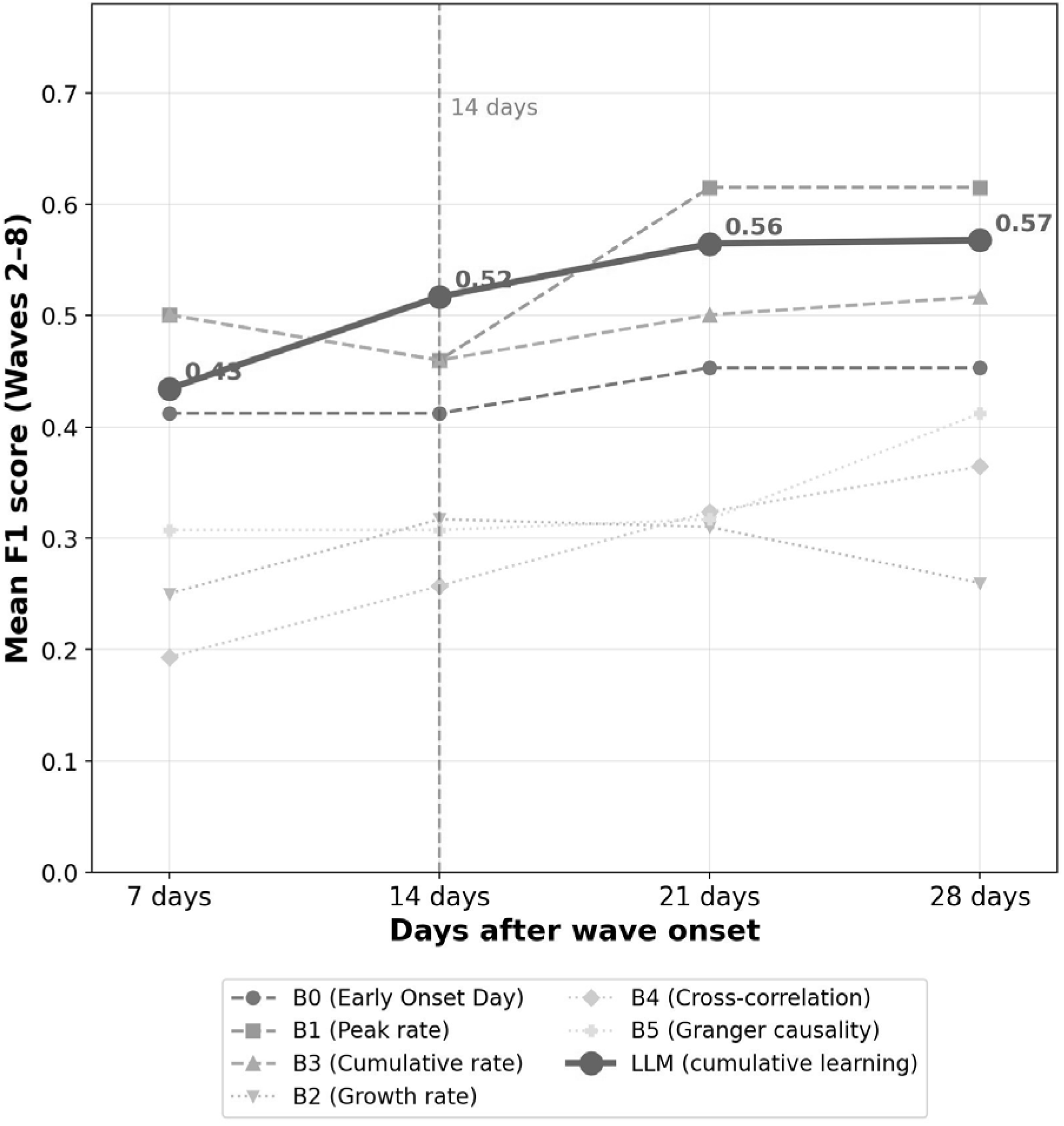
Mean F1 scores by method across time points after wave onset (Waves 2–8) Mean F1 scores (Waves 2–8) at 7, 14, 21, and 28 days after wave onset for the cumulative-learning LLM (blue solid line) and six non-cumulative statistical baselines (B0–B5, grey dashed/dotted lines). The vertical dashed red line marks the 14-day reference time point. Numerical F1 values for the LLM are annotated at each time point. All baseline methods are non-cumulative; the LLM uses cumulative historical context from all prior waves. Wave 6 (Omicron BA.1) is excluded from mean calculations as it was undetectable by all methods (F1 = 0.00). *B0: Early Onset Day; B1: peak infection rate; B2: OLS growth rate; B3: cumulative infection rate; B4: cross-correlation lead score; B5: Granger causality score. LLM: cumulative-learning LLM (Claude Haiku, claude-haiku-4-5-20251001)*.

**Figure 3.**
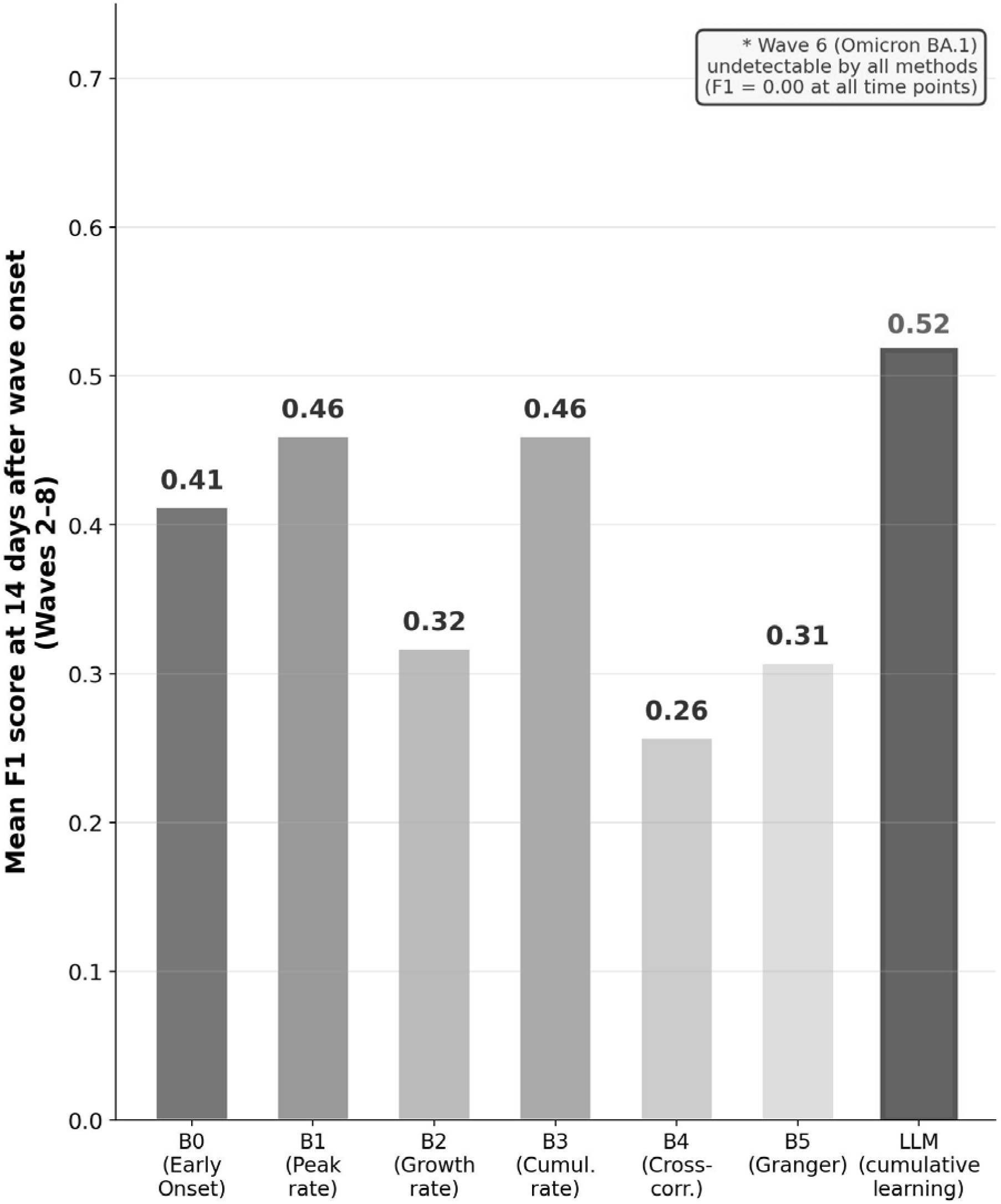
Mean F1 scores at 14 days after wave onset by method (Waves 2–8) Bar chart comparing mean F1 scores at 14 days after wave onset across six non-cumulative statistical baselines (B0–B5, grey bars) and the cumulative-learning LLM (blue bar). Numerical F1 values are annotated above each bar. The box annotation indicates that Wave 6 (Omicron BA.1) was undetectable by all methods (F1 = 0.00 at all time points) and is included in the mean calculations. *B0: Early Onset Day (0.41); B1: Peak infection rate (0.46); B2: OLS growth rate (0.32); B3: Cumulative infection rate (0.46); B4: Cross-correlation lead score (0.26); B5: Granger causality score (0.31); LLM: cumulative-learning LLM (0.52)*.

### 3.2 Cumulative Calculation: Statistical Baselines

Cumulative calculation versions of B1 and B3 achieved mean F1 = 0.51 at 14 days, closely matching the LLM (F1 = 0.52; Table 1, Figure 2). Wave 7 achieved F1 = 1.00 across all cumulative methods; Wave 6 remained undetected (F1 = 0.00), confirming that the detection failure is attributable to input data rather than analytical method. The λ = 0 condition consistently yielded lower performance, confirming that historical origin weighting contributes meaningfully beyond current-wave signals alone (Table S1, Table S2).

**Table 1.** Mean F1 scores by method and time point after wave onset (Waves 2–8)

| Method | 7 days | 14 days | 21 days | 28 days |
| --- | --- | --- | --- | --- |
| B0 (Early Onset Day) | 0.41 | 0.41 | 0.45 | 0.45 |
| B1 (Peak infection rate) | 0.50 | 0.46 | 0.61 | 0.61 |
| B2 (OLS growth rate) | 0.25 | 0.32 | 0.31 | 0.26 |
| B3 (Cumulative rate) | 0.50 | 0.46 | 0.50 | 0.52 |
| B4 (Cross-correlation) | 0.19 | 0.26 | 0.32 | 0.36 |
| B5 (Granger causality) | 0.31 | 0.31 | 0.32 | 0.41 |
| LLM (non-cumulative) | 0.46 | 0.46 | 0.61 | 0.57 |
| LLM (cumulative) | 0.44 | 0.52 | 0.57 | 0.57 |

*F1 scores against genomically confirmed regional wave origins (11-block system). B0–B5: non-cumulative statistical baselines (each wave treated independently). LLM (non-cumulative): LLM with current-wave data only, no historical context. LLM (cumulative): cumulative-learning LLM incorporating confirmed origins from all prior waves. Wave 6 (Omicron BA.1) was undetectable by all methods ( $F1 = 0.00$ at all time points) and is included in the mean.*

**Table 2.**
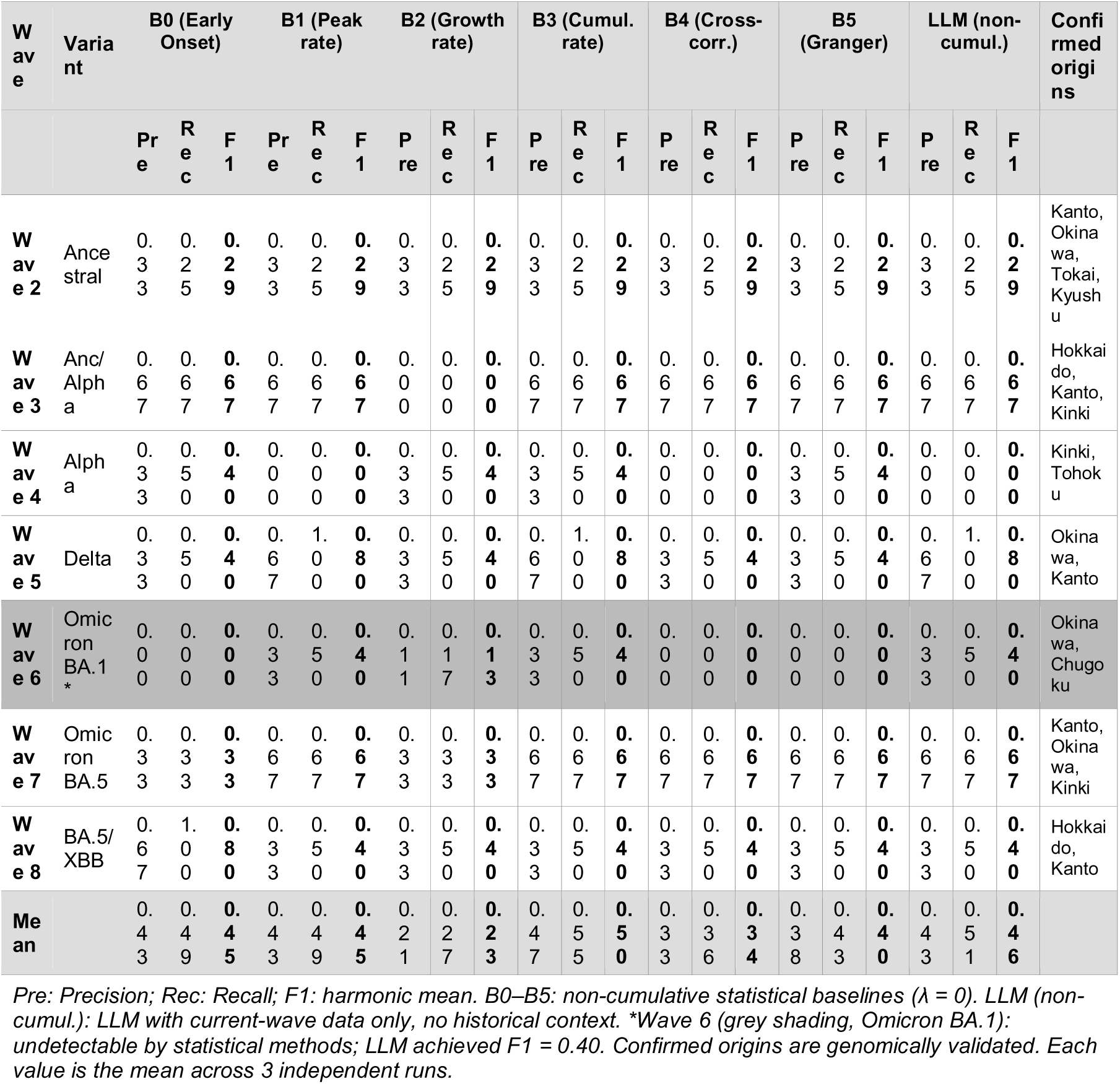
Wave-specific Precision, Recall, and F1 at 14 days: non-cumulative statistical baselines (Waves 2–8)

### 3.3 Cumulative Learning: LLM Performance

The cumulative-learning LLM achieved mean F1 = 0.44 (7 days), 0.52 (14 days), 0.57 (21 and 28 days), exceeding all non-cumulative statistical baselines at 14 days (Figure 2, Figure 3). Wave 7 achieved F1 = 1.00 at 14 days; Wave 5 (Delta) was detected from day 7 (F1 = 0.80). Wave 6 was undetected at all time points (F1 = 0.00). Outputs were stable across three independent runs (Figure 4).

**Figure 4.**
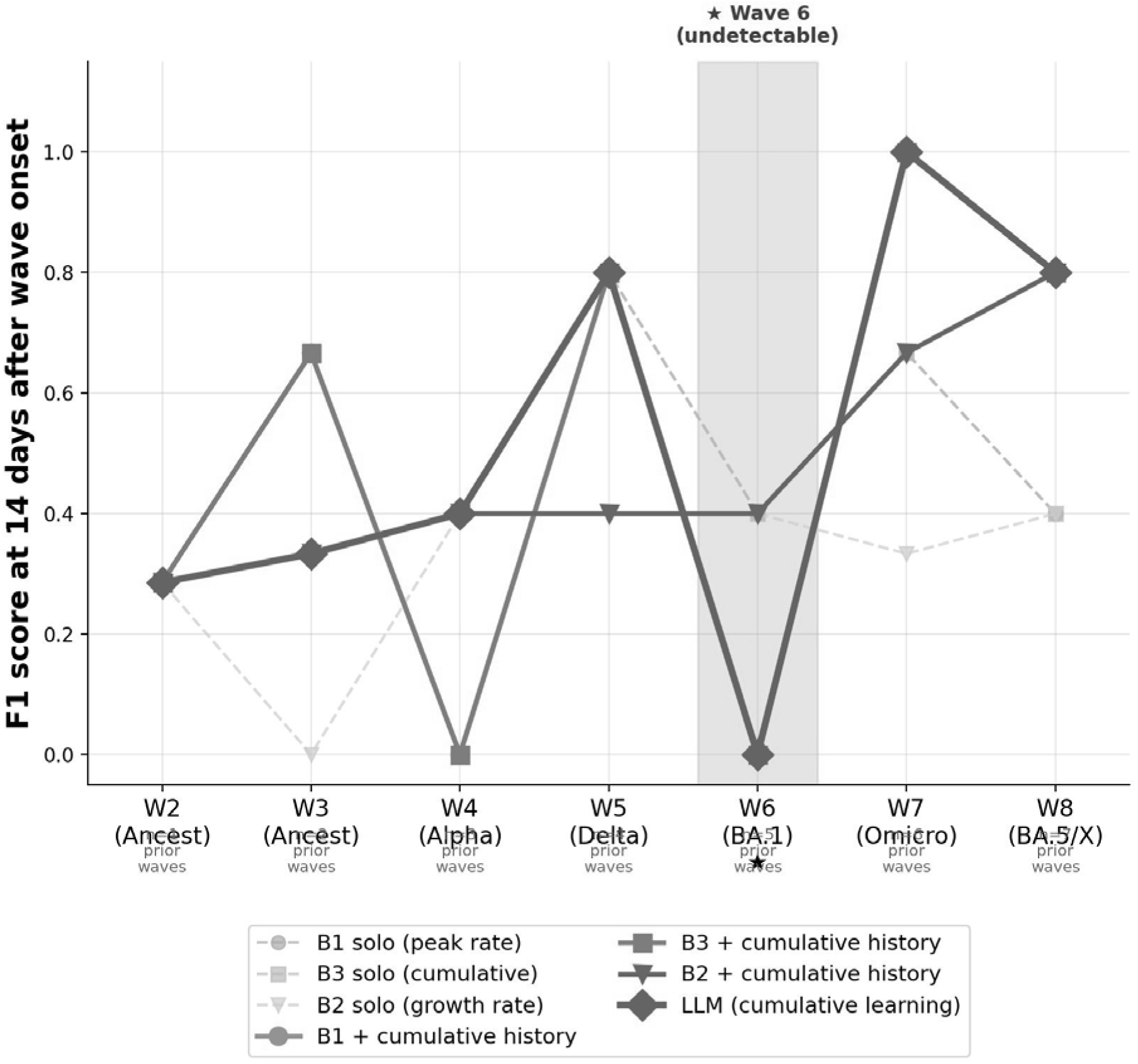
Wave-specific F1 scores at 14 days after wave onset: cumulative methods vs. non-cumulative baselines. Wave-specific F1 scores at 14 days after wave onset for the cumulative-learning LLM (blue diamond), cumulative calculation versions of B1, B2, and B3 (colored lines), and corresponding non-cumulative baselines (grey dashed lines). The x-axis shows each wave with its dominant variant and the number of prior waves available for cumulative weighting. Wave 6 (Omicron BA.1) is highlighted in red shading to indicate that it was undetectable by all methods (F1 = 0.00). Convergence of cumulative LLM and cumulative statistical methods in Waves 7 and 8 illustrates the equivalence of the two approaches. *LLM: cumulative-learning LLM; B1+hist: cumulative calculation B1 (*λ*=0.75); B2+hist: cumulative calculation B2 (*λ*=0.75); B3+hist: cumulative calculation B3 (*λ*=0.75). Solo variants (B1/B2/B3 solo) represent non-cumulative baselines for comparison*.

### 3.4 Combination of Statistical Indicators with LLM

Providing statistical indicator outputs as additional input to the LLM did not consistently improve performance. The LLM appeared to override noisy statistical inputs and rely primarily on cumulative historical context. Details are provided in Table 3 and Table 4.

**Table 3.**
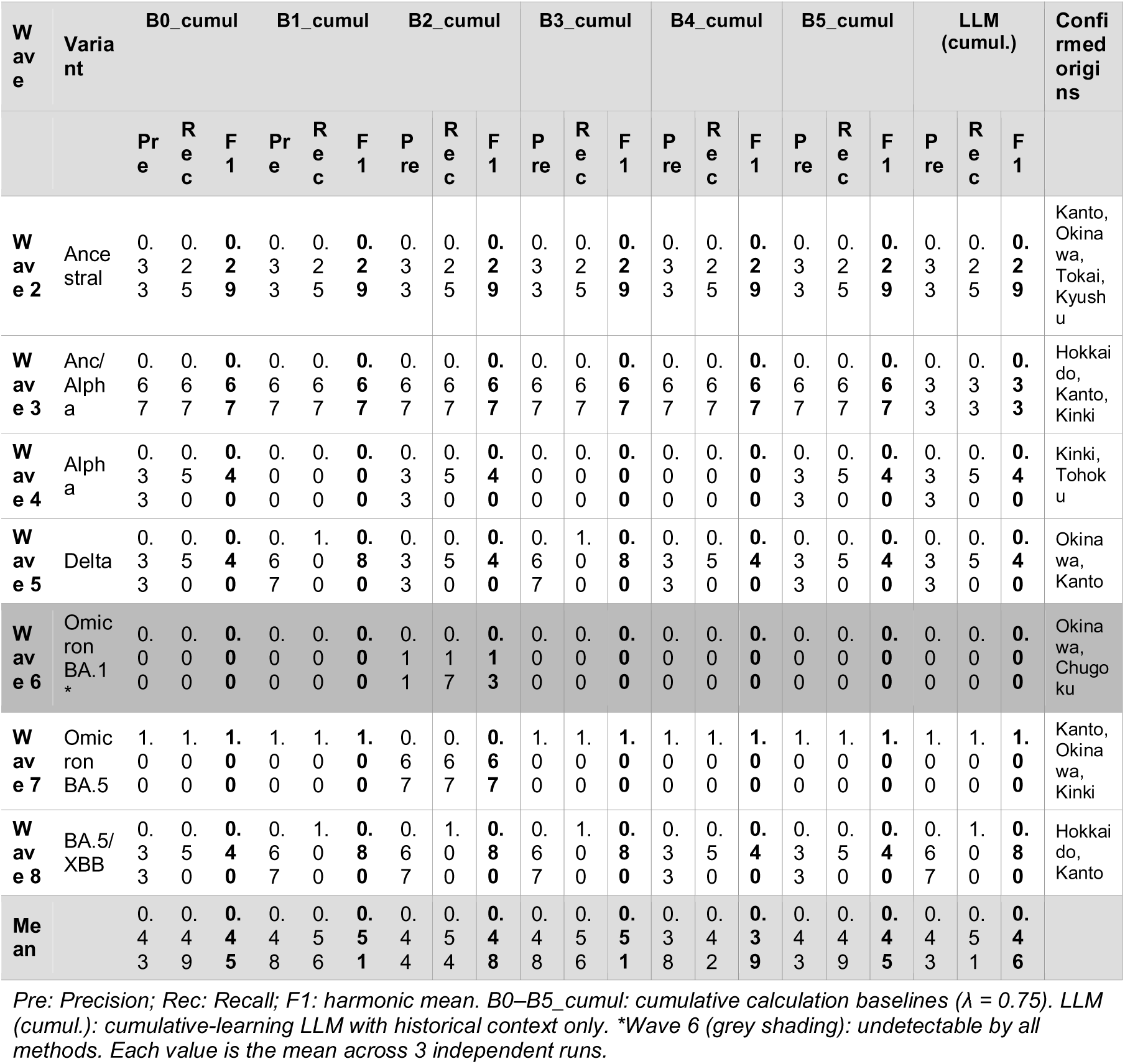
Wave-specific Precision, Recall, and F1 at 14 days: cumulative calculation baselines and cumulative-learning LLM (Waves 2–8)

**Table 4.** Wave-specific Precision, Recall, and F1 at 14 days: combined cumulative statistical × LLM methods (Waves 2–8)

| Wave | Variant | B0+LLM |  |  | B1+LLM |  |  | B2+LLM |  |  | B3+LLM |  |  | B4+LLM |  |  | B5+LLM |  |  | Confirmed origins |
| --- | --- | --- | --- | --- | --- | --- | --- | --- | --- | --- | --- | --- | --- | --- | --- | --- | --- | --- | --- | --- |
|  |  | Pre | Rec | F1 | Pre | Rec | F1 | Pre | Rec | F1 | Pre | Rec | F1 | Pre | Rec | F1 | Pre | Rec | F1 |  |
| Wave 2 | Ancestral | 0.33 | 0.25 | **0.29** | 0.33 | 0.25 | **0.29** | 0.33 | 0.25 | **0.29** | 0.33 | 0.25 | **0.29** | 0.33 | 0.25 | **0.29** | 0.33 | 0.25 | **0.29** | Kanto, Okinawa, Tokai, Kyushu |
| Wave 3 | Anc/Alpha | 0.33 | 0.33 | **0.33** | 0.67 | 0.67 | **0.67** | 1.00 | 1.00 | **1.00** | 0.67 | 0.67 | **0.67** | 0.67 | 0.67 | **0.67** | 0.67 | 0.67 | **0.67** | Hokkaido, Kanto, Kinki |
| Wave 4 | Alpha | 0.33 | 0.50 | **0.40** | 0.00 | 0.00 | **0.00** | 0.33 | 0.50 | **0.40** | 0.33 | 0.50 | **0.40** | 0.33 | 0.50 | **0.40** | 0.33 | 0.50 | **0.40** | Kinki, Tohoku |
| Wave 5 | Delta | 0.33 | 0.50 | **0.40** | 0.67 | 1.00 | **0.80** | 0.33 | 0.50 | **0.40** | 0.67 | 1.00 | **0.80** | 0.33 | 0.50 | **0.40** | 0.33 | 0.50 | **0.40** | Okinawa, Kanto |
| Wave 6 | Omicron BA.1* | 0.00 | 0.00 | **0.00** | 0.00 | 0.00 | **0.00** | 0.33 | 0.50 | **0.40** | 0.00 | 0.00 | **0.00** | 0.00 | 0.00 | **0.00** | 0.00 | 0.00 | **0.00** | Okinawa, Chugoku |
| Wave 7 | Omicron BA.5 | 1.00 | 1.00 | **1.00** | 1.00 | 1.00 | **1.00** | 0.67 | 0.67 | **0.67** | 1.00 | 1.00 | **1.00** | 1.00 | 1.00 | **1.00** | 1.00 | 1.00 | **1.00** | Kanto, Okinawa, Kinki |
| Wave 8 | BA.5/XBB | 0.67 | 1.00 | **0.80** | 0.67 | 1.00 | **0.80** | 0.67 | 1.00 | **0.80** | 0.67 | 1.00 | **0.80** | 0.33 | 0.50 | **0.40** | 0.67 | 1.00 | **0.80** | Hokkaido, Kanto |
| Mean |  | 0.43 | 0.51 | **0.46** | 0.48 | 0.56 | **0.51** | 0.52 | 0.63 | **0.57** | 0.52 | 0.63 | **0.57** | 0.43 | 0.49 | **0.45** | 0.48 | 0.56 | **0.51** |  |

Pre: Precision; Rec: Recall; F1: harmonic mean. B0–B5+LLM: cumulative statistical indicator ( $\lambda = 0.75$ ) with prompt-guided LLM weighting. \*Wave 6 (grey shading): undetectable by all methods. Each value is the mean across 3 independent runs.

### 3.5 Wave 6 Exception

Wave 6 (Omicron BA.1, November 2021–June 2022) was undetected by all six statistical baselines, all cumulative statistical methods, and the cumulative-learning LLM across all time points (F1 = 0.00). The genomically confirmed origins—Okinawa and Chugoku—correspond to regions with documented early cluster events that preceded entry into the general domestic surveillance system. This systematic failure across all methods and all time windows is consistent with an introduction pathway that was not captured in routine prefecture-level case count data. The specific mechanism underlying this surveillance gap remains a subject for future investigation.

## 4. Discussion

### 4.1 Principal Findings

This study demonstrates three principal findings. First, a general-purpose LLM applied without fine-tuning, using only routinely available case count data, outperformed conventional statistical methods for early identification of COVID-19 wave origins in Japan. Second, the cumulative-learning principle underlying this advantage—weighting regions that have historically served as wave origins—can be replicated using simple, transparent arithmetic applied to the same statistical baselines. Third, the convergence of LLM and cumulative statistical methods in later waves (Waves 7 and 8) suggests that the key mechanism is historical origin weighting rather than LLM-specific reasoning or pattern recognition.

The performance advantage was most pronounced at 14 days after wave onset (LLM F1 = 0.52 vs. 0.41–0.46 for the best static baselines), a clinically relevant time window for public health decision-making. The strong performance in Wave 5 (Delta, F1 = 0.80 from day 7) and Wave 7 (Omicron BA.5, F1 = 1.00 at day 14) suggests that the cumulative approach successfully captures the relationship between regional immunity status from prior waves and subsequent wave origin locations. In both of these waves, Okinawa was correctly identified as an origin region from early time points—consistent with its documented role as an international gateway and its repeatedly confirmed status as an early-wave origin across multiple prior waves. The cumulative learning mechanism effectively encodes this historical pattern and applies it as a prior for subsequent wave prediction. The underperformance of B2 (OLS growth rate), B4 (cross-correlation), and B5 (Granger causality) relative to the simpler B1 and B3 baselines is noteworthy. These more sophisticated statistical approaches rely on temporal dynamics computed over the first N days, which are inherently noisy in the early phase of a wave when case counts are low. In contrast, B1 (peak infection rate) and B3 (cumulative infection rate) are more robust to noise because they summarize the overall level of infection rather than the rate of change. This finding suggests that for very early detection (7–14 days), simpler magnitude-based metrics may outperform more complex dynamic metrics. The non-cumulative LLM—operating without any historical context—achieved mean F1 = 0.46 at 14 days, only 0.06 below the cumulative version (F1 = 0.52). This small gap suggests that the LLM possesses substantial intrinsic geographic reasoning capacity that operates independently of accumulated historical knowledge. Particularly striking is the non-cumulative LLM’s detection of Wave 6 (F1 = 0.40), which all statistical methods—cumulative or otherwise—failed to identify (F1 = 0.00). This indicates that LLM reasoning may access epidemiological priors embedded in training data (e.g., knowledge of Okinawa’s role as an international gateway) that statistical methods cannot exploit. The marginal performance advantage of the cumulative LLM over the non-cumulative LLM (F1 = 0.52 vs. 0.46 at 14 days) is modest, while the cumulative statistical baselines match the cumulative LLM (F1 = 0.51 vs. 0.52). This suggests that the key mechanism driving cumulative improvement is historical origin weighting—which can be implemented in a spreadsheet—rather than the LLM’s contextual reasoning per se. This result suggests that LLMs do not universally outperform simpler approaches for structured spatial reasoning tasks in epidemiology. Researchers and practitioners considering LLM deployment in surveillance settings are encouraged to benchmark against transparent statistical baselines before concluding that AI-based approaches offer meaningful advantages. Future work should address not only prompt engineering refinements but also the fundamental algorithmic capacity of LLMs to integrate epidemiological pattern recognition with geographic reasoning—until such advances are demonstrated, cumulative calculation methods remain the recommended first-line approach. It should also be noted that the LLM did not process raw surveillance data directly; rather, it received human-designed epidemiological summaries—ranked regional incidence, Early Onset Day values, and historical origin patterns structured by the investigators. A portion of the observed LLM performance may therefore reflect the quality of this feature engineering rather than intrinsic LLM reasoning capacity, and performance gains from prompt refinement may be difficult to disentangle from improvements in feature design. It should also be noted that the LLM did not process raw surveillance data directly; rather, it received human-designed epidemiological summaries—ranked regional incidence, Early Onset Day values, and historical origin patterns structured by the investigators. A portion of the observed LLM performance may therefore reflect the quality of this feature engineering rather than intrinsic LLM reasoning capacity, and performance gains from prompt refinement may be difficult to disentangle from improvements in feature design.

### 4.2 Transparency and Interpretability

The cumulative calculation framework was conceptually inspired by Bayesian updating: P(r,n) functions as an empirical prior—the historical tendency of a region to serve as a wave origin—while Score_baseline(r) serves as the likelihood term reflecting current-wave evidence. Yet the method deliberately avoids formal Bayesian specification, requiring no distributional assumptions, no posterior computation, and no statistical software beyond a spreadsheet. This simplification proved sufficient to match LLM performance, suggesting that the core signal lies in the historical weighting mechanism itself. The finding that cumulative calculation statistical baselines (B1_cumul, B3_cumul) achieved performance closely matching the LLM has important practical implications. Unlike the LLM, which processes historical context through a black-box neural network, the cumulative statistical methods make every weighting decision fully explicit and auditable: the past origin score P(r,n) is simply the proportion of prior waves in which region r was a confirmed origin, and the final ranking is a straightforward linear combination of this score and a normalized current-wave indicator. This transparency facilitates interpretation, regulatory review, and adaptation to local contexts without requiring access to proprietary model APIs.

The convergence of LLM and cumulative statistical predictions in Waves 7 and 8 further supports the interpretation that both approaches are leveraging the same underlying signal—the historical tendency of certain regions (Kanto, Okinawa, Kinki) to serve as wave origins—rather than the LLM extracting qualitatively different epidemiological insight from the data. From a practical deployment perspective, the transparent statistical implementation offers several operational advantages. It can be implemented in any spreadsheet or scripting environment, requires no internet connection or API dependency, can be audited line-by-line by public health officials, and produces results that are directly explainable to decision-makers without reference to machine learning concepts. For settings where LLM API access is unavailable or where regulatory requirements mandate explainable algorithms, the cumulative statistical approach provides a deployable alternative that sacrifices minimal performance relative to the LLM (mean F1 = 0.51 vs. 0.52 at 14 days).

### 4.3 Wave 6 and Surveillance Data Limitations

The uniform failure of all methods in Wave 6 constitutes an important negative finding with direct public health implications. The confirmed origins of the Omicron BA.1 wave in Okinawa and Chugoku involved early cluster events that preceded entry into the standard prefectural reporting pipeline and were therefore absent from the case count data used by all methods in this study. This finding illustrates a structural limitation of domestic case surveillance data for origin identification: when introduction events occur through channels not covered by routine surveillance, neither advanced statistical methods nor cumulative-learning LLMs can compensate for the absence of signal. Integration of genomic surveillance data, international travel records, and enhanced local outbreak reporting would likely be necessary to address this gap.

### 4.4 Implications for Epidemic Surveillance and Public Health Practice

These results suggest that cumulative-learning approaches—whether implemented via LLM or transparent statistical methods—may serve as practical, low-cost complements to conventional surveillance analysis. Both can be applied immediately when a new wave begins, requiring only routinely available case count data and confirmed origins from prior waves, without waiting for genomic results or specialized laboratory infrastructure. The 14-day time window for achieving mean F1 = 0.52 has direct operational relevance: in the Japanese context, 14 days after wave onset corresponds to the period during which case counts are rising but have not yet reached regional peak—precisely the window in which targeted early intervention, such as enhanced contact tracing in candidate origin regions or travel advisories, could meaningfully slow geographic spread. The system’s output—a ranked list of three candidate origin regions—is directly actionable, providing a probabilistic shortlist to guide prioritization of surveillance resources rather than requiring the certainty that genomic surveillance itself rarely provides in real time. The failure of all methods in Wave 6 (Omicron BA.1) serves as an important calibration point: when a wave originates through channels not captured by routine surveillance, no analytical method can compensate for the absence of signal, and explicit data quality monitoring would be required in operational deployment.

Beyond COVID-19, the framework is data-agnostic and applicable to any surveillance system providing (1) a regional incidence time series, (2) a defined aggregation scheme, and (3) a historical record of confirmed wave origins. Japan’s Infectious Disease Weekly Report (IDWR) sentinel surveillance, for example, provides weekly influenza incidence at the prefecture level spanning more than a decade—sufficient historical depth to deploy the same cumulative-learning framework for real-time influenza wave origin estimation at no additional data collection cost. The results should not be interpreted as evidence that these approaches are ready to replace established epidemiological methods: F1 scores of 0.51–0.52 indicate that approximately half of predicted origins are incorrect or confirmed origins are missed. The approach is best understood as a hypothesis-generating complement to genomic surveillance, not a replacement.

### 4.5 Limitations

Several limitations warrant consideration. First, the evaluation was conducted on eight epidemic waves in a single country; confidence intervals are wide and results are best interpreted as exploratory rather than confirmatory. Second, genomic validation data have varying evidence quality across waves—Wave 6 origins in particular lack independent NIID peer-reviewed confirmation. Third, the LLM is a proprietary model whose internal representations are not interpretable; whether performance reflects genuine epidemiological reasoning or pattern-matching with training data cannot be determined. Fourth, all analyses are retrospective, and prospective validation across different countries, pathogens, and surveillance systems is essential before operational deployment. Design parameters—including the detection threshold (0.1/100,000), the top-3 prediction rule, and λ—were set without systematic optimization and may require recalibration in other settings. Future priorities include formal λ optimization, alternative weighting schemes (temporal decay, variant-specific weighting), and integration of additional data sources such as mobility networks and wastewater surveillance. A leave-one-wave-out analysis of λ selection confirmed that λ ≥ 0.25 was optimal in 6 of 7 waves, supporting the robustness of the chosen λ = 0.75 (Table S1, Table S2). The exception was Wave 8 (BA.5/XBB), suggesting that cumulative weighting may be less beneficial when successive waves share the same variant lineage—a finding that motivates adaptive λ selection as a direction for future work. A leave-one-wave-out analysis of λ selection confirmed that λ ≥ 0.25 was optimal in 6 of 7 waves, supporting the robustness of the chosen λ = 0.75 (Table S1, Table S2). The exception was Wave 8 (BA.5/XBB), suggesting that cumulative weighting may be less beneficial when successive waves share the same variant lineage—a finding that motivates adaptive λ selection as a direction for future work.

## 5. Conclusions

This study demonstrates that simple cumulative calculation of historical origin frequencies performs comparably to a cumulative-learning LLM for real-time geographic origin identification of ongoing COVID-19 waves (mean F1 = 0.51 vs. 0.52 at 14 days after wave onset). The marginal advantage of 0.01 in F1 does not justify the additional cost and complexity of LLM deployment in routine public health practice. We provide the complete four-step formulation of the cumulative calculation method (Box 1) as a ready-to-implement tool requiring only a spreadsheet and routinely available surveillance data. These findings suggest that LLMs do not universally outperform simpler approaches, and that the key mechanism is historical origin weighting—not AI reasoning. Future work should determine whether prompt engineering advances or algorithmic improvements can push LLM performance substantially above the cumulative statistical ceiling before LLM-specific deployment is warranted. The systematic failure of all methods in Wave 6 underscores that no analytical approach can compensate for absent surveillance signals; the method is best understood as a hypothesis-generating complement to genomic surveillance, not a replacement.

## Declarations

### Ethics approval and consent to participate

This study used publicly available aggregate surveillance data. No individual patient data were used. Ethics approval was not required.

### Consent for publication

Not applicable.

### Availability of data and materials

Prefecture-level COVID-19 case data are available from the MHLW open data portal (https://www.mhlw.go.jp/stf/covid-19/open-data.html). Analysis code will be made available on GitHub upon acceptance.

### Conflict of Interest

The authors declare that they have no conflict of interest.

### Funding

No external funding was received for this study. The authors declare that no funding sources influenced study design, data collection, analysis, or interpretation.

### Authors’ contributions

SN conceived the study, developed the methodology, conducted all analyses, and drafted the manuscript. AY contributed to study design, interpretation of results, and critical revision of the manuscript. Both authors approved the final version.

## Data Availability

Prefecture-level COVID-19 case data are available from the Ministry of Health, Labour and Welfare open data portal (https://www.mhlw.go.jp/stf/covid-19/open-data.html). Analysis code will be made available on GitHub upon acceptance.

## Acknowledgements

The authors thank the Ministry of Health, Labour and Welfare of Japan for making prefecture-level COVID-19 surveillance data publicly available. Special thanks to Dr. Hiroaki Kitano (Okinawa Institute of Science and Technology, OIST) and Prof. Akimasa Hirata (Nagoya Institute of Technology) for their insightful advice. The authors used Claude (Anthropic, claude.ai), an LLM-based AI assistant, to support coding, data analysis, and manuscript preparation. All results and conclusions were verified by the authors.

## AI Use Disclosure

The authors used Claude (Anthropic, claude.ai), an LLM-based AI assistant, to support data analysis, drafting, figure generation, and coding. All results and conclusions were verified by the authors.

## Supplementary Material

This supplementary material provides sensitivity analyses for the hyperparameter λ used in the cumulative calculation method.

**Table S1.** Mean F1 scores by cumulative calculation baseline and λ value (14 days after wave onset, Waves 2–8)

| Method | $\lambda = 0.00$ | $\lambda = 0.25$ | $\lambda = 0.50$ | $\lambda = 0.75$ | $\lambda = 1.00$ |
| --- | --- | --- | --- | --- | --- |
| B0_cumul | 0.41 | **0.45** | **0.45** | **0.45** | **0.45** |
| B1_cumul | 0.46 | **0.51** | **0.51** | **0.51** | **0.51** |
| B2_cumul | 0.28 | **0.41** | **0.47** | **0.52** | **0.52** |
| B3_cumul | 0.46 | **0.45** | **0.51** | **0.51** | **0.51** |

Mean F1 scores at 14 days after wave onset across Waves 2–8. Bold values indicate improvement over the non-cumulative baseline ( $\lambda = 0.00$ ). B0\_cumul shows no $\lambda$ -dependence due to the discrete nature of onset-day ranking. B1 and B3 stabilize at $\lambda \geq 0.25$ ; B2 peaks at $\lambda = 0.75$ .

**Table S2.** Leave-one-wave-out λ robustness analysis for B1 (14 days after wave onset)

| Held-out Wave | Remaining 6-wave F1 ($\lambda=0.00$) | Remaining 6-wave F1 ($\lambda=0.75$) | Optimal $\lambda$ | Held-out F1 ($\lambda=0.75$) |
| --- | --- | --- | --- | --- |
| Wave 2 | 0.489 | 0.545 | $\geq 0.25$ | 0.286 |
| Wave 3 | 0.426 | 0.481 | $\geq 0.25$ | 0.667 |
| Wave 4 | 0.537 | 0.592 | $\geq 0.25$ | 0.000 |
| Wave 5 | 0.403 | 0.459 | $\geq 0.25$ | 0.800 |
| Wave 6 | 0.470 | 0.592 | $\geq 0.25$ | 0.000 |
| Wave 7 | 0.425 | 0.425 | $\geq 0.25$ | 1.000 |
| Wave 8 | 0.470 | 0.459 | 0.00 | 0.800 |

For each held-out wave, the remaining six waves were used to compare $\lambda = 0.00$ versus $\lambda = 0.75$ for B1\_cumul. Optimal $\lambda$ was selected based on the remaining six waves; the held-out wave's F1 at $\lambda = 0.75$ is reported for reference. In 6 of 7 waves, $\lambda \geq 0.25$ was optimal, confirming robustness of the $\lambda = 0.75$ selection. The exception was Wave 8 (BA.5/XBB), where successive same-lineage waves may reduce the predictive value of historical weighting.

## Notes

### Competing Interest Statement

The authors have declared no competing interest.

### Author Declarations

The study used ONLY openly available human data that were originally located at: the Ministry of Health, Labour and Welfare (MHLW) open data portal (https://www.mhlw.go.jp/stf/covid-19/open-data.html).

## References

[1] Kraemer MUG, et al. The effect of human mobility and control measures on the COVID-19 epidemic in China. Science. 2020;368(6490):493–497.

[2] Hadfield J, et al. Nextstrain: real-time tracking of pathogen evolution. Bioinformatics. 2018;34(23):4121–4123.

[3] Granger CWJ. Investigating causal relations by econometric models and cross-spectral methods. Econometrica. 1969;37(3):424–438.

[4] Wallinga J, Lipsitch M. How generation intervals shape the relationship between growth rates and reproductive numbers. Proc Biol Sci. 2007;274(1609):599–604.

[5] Chowell G, et al. Characterizing the reproduction number of epidemics with early subexponential growth dynamics. J R Soc Interface. 2016;13(123):20160659.

[6] Du H, Zhao J, Zhao Y, et al. Advancing real-time pandemic forecasting using large language models: A COVID-19 case study. arXiv:2404.06962. 2024.

[7] Deleted.

[8] Jin M, et al. Time-LLM: Time Series Forecasting by Reprogramming Large Language Models. arXiv:2310.01728. 2023.

[9] Li J, Lai S, Gao GF, Shi W. The emergence, genomic diversity and global spread of SARS-CoV-2. Nature. 2021;600(7889):408–418.

[10] Rambaut A, Holmes EC, O’Toole A, et al. A dynamic nomenclature proposal for SARS-CoV-2 lineages to assist genomic epidemiology. Nat Microbiol. 2020;5(11):1403–1407.

[11] Hirotsu Y, Omata M. SARS-CoV-2 Omicron sublineage BA.2 replaces BA.1.1: genomic surveillance in Japan from September 2021 to March 2022. J Infect. 2022;85(1):e20-e22.

[12] Sasanami M, Kayano T, Nishiura H. Monitoring the COVID-19 immune landscape in Japan. Int J Infect Dis. 2022;122:300–306.

[13] Ito K, Piantham C, Nishiura H. Predicted dominance of variant Delta of SARS-CoV-2 before Tokyo Olympic Games, Japan, July 2021. Euro Surveill. 2021;26(27):2100570.

[14] Kwok KO, Huynh T, Wei WI, Wong SYS, Riley S, Tang A. Utilizing large language models in infectious disease transmission modelling for public health preparedness. Comput Struct Biotechnol J. 2024;23:3254–3257.

[15] Cascella M, Sepe R, Napolitano F, et al. Utilizing natural language processing and large language models in the diagnosis and prediction of infectious diseases: a systematic review. Am J Infect Control. 2024;52(7):825–834.

[16] McMahon T, Chan A, Havlin S, Gallos LK. Spatial correlations in geographical spreading of COVID-19 in the United States. Sci Rep. 2022;12(1):699.

[17] Zhao Y, Xu X, Shen M, Du Z, Yang W, Feng L. Revealing associations between spatial time series trends of COVID-19 incidence and human mobility. Epidemiol Infect. 2023;151:e198.

[18] Furuse Y, Sando E, Tsuchiya N, et al. Clusters of coronavirus disease in communities, Japan, January-April 2020. Emerg Infect Dis. 2020;26(9):2176–2179.

[19] Nishiura H, Linton NM, Akhmetzhanov AR. Serial interval of novel coronavirus (COVID-19) infections. Int J Infect Dis. 2020;93:284–286.

[20] Morens DM, Fauci AS. Emerging pandemic diseases: how we got to COVID-19. Cell. 2020;182(5):1077–1092.

[21] Tegally H, Moir M, Everatt J, et al. Dispersal patterns and influence of air travel during the global expansion of SARS-CoV-2 variants of concern. Cell. 2023;186(15):3277–3290.

[22] Tanaka H, Ogata T, Shibata T, et al. Shorter incubation period among COVID-19 cases with the BA.1 Omicron variant. Int J Environ Res Public Health. 2022;19(10):6330.

[23] Hayashi K, Kayano T, Anzai A, et al. Assessing public health and social measures against COVID-19 in Japan from March to June 2021. Front Med (Lausanne). 2022;9:937732.

[24] Lu Y, Cai G, Hu Z, et al. Exploring spatiotemporal patterns of COVID-19 infection in Nagasaki Prefecture in Japan. Arch Public Health. 2022;80(1):177.

[25] Cowling BJ, Ali ST, Ng TWY, et al. Impact assessment of non-pharmaceutical interventions against coronavirus disease 2019 and influenza in Hong Kong. Lancet Public Health. 2020;5(5):e279–e288.

[26] Flaxman S, Mishra S, Gandy A, et al. Estimating the effects of non-pharmaceutical interventions on COVID-19 in Europe. Nature. 2020;584(7820):257–261.

[27] Vespignani A, Tian H, Dye C, et al. Modelling COVID-19. Nat Rev Phys. 2020;2(6):279–281.

[28] Volz E, Hill V, McCrone JT, et al. Evaluating the effects of SARS-CoV-2 spike mutation D614G on transmissibility and pathogenicity. Cell. 2021;184(1):64–75.

[29] Mossong J, Hens N, Jit M, et al. Social contacts and mixing patterns relevant to the spread of infectious diseases. PLoS Med. 2008;5(3):e74.

[30] Tuite AR, Bogoch II, Sherbo R, Watts A, Fisman D, Khan K. Estimation of coronavirus disease 2019 (COVID-19) burden and potential for international dissemination of infection from Iran. Ann Intern Med. 2020;172(10):699–701.

[31] Brown JR, Goldstein RA, Bhatt S, et al. Predicting the epidemic kinetics of SARS-CoV-2 from sequence data in real time. Nat Commun. 2022;13(1):4393.

[32] Nakagawa S, Yamamoto A. Relationship between the effective reproduction number and cumulative incidence of prior waves: analysis of eight COVID-19 epidemic waves in Japan. Environ Health Prev Med 2025. [In press]

[33] Sasanami M, Kayano T, Nishiura H. Wave-specific estimation of COVID-19 infection fatality risk in Japan. J Theor Biol. 2023;559:111384.

[34] Nishiura H, Kobayashi T, Miyama T, et al. Estimation of the asymptomatic ratio of novel coronavirus infections (COVID-19). Int J Infect Dis. 2020;94:154–155.

[35] Keeling MJ, Woolhouse MEJ, Shaw DJ, et al. Dynamics of the 2001 UK foot and mouth epidemic: stochastic dispersal in a heterogeneous landscape. Science. 2001;294(5543):813–817.

